# Public attitudes towards the use of novel technologies in their future healthcare: A UK survey

**DOI:** 10.1101/2021.12.05.21266892

**Authors:** Sarah Sauchelli, Tim Pickles, Alexandra Voinescu, Heungjae Choi, Ben Sherlock, Jingjing Zhang, Steffi Colyer, Sabrina Grant, Sethu Sundari, Gemma Lasseter

## Abstract

**Background:** Innovation in healthcare technologies can result in more convenient and effective treatment that is less costly, but a persistent challenge to widespread adoption in health and social care is end user acceptability. The purpose of this study was to capture UK public opinions and attitudes to novel healthcare technologies (NHTs), and to better understand the factors that contribute to acceptance and future use.

**Methods:** An online survey was distributed to the UK public between April and May 2020. Respondents received brief information about four novel healthcare technologies (NHTs) in development: a laser-based tool for early diagnosis of osteoarthritis, a virtual reality tool to support diabetes self-management, a non-invasive continuous glucose monitor using microwave signals, a mobile app for patient reported monitoring of rheumatoid arthritis. They were queried on their general familiarity and attitudes to technology, and their willingness to accept each NHT in their future care. Responses were analysed using summary statistics and content analysis.

**Results:** Knowledge about NHTs was diverse, with respondents being more aware about the health applications of mobile apps (66%), followed by laser-based technology (63.8%), microwave signalling (28%), and virtual reality (18.3%). Increasing age and the presence of a self-reported medical condition favoured acceptability for some NHTs, whereas self-reported understanding of how the NHT works resulted in elevated acceptance scores across all NHTs presented. Common contributors to hesitancy were safety and risks from use. Respondents wanted more information and evidence to help inform their decisions, ideally provided verbally by a general practitioner or health professional. Other concerns, such as privacy, were NHT-specific but equally important in decision-making.

**Conclusions:** Early insight into the knowledge and preconceptions of the public about NHTs in development can assist their design and prospectively mitigate obstacles to acceptance and adoption.

## Background

In the United Kingdom there are approximately 12 million older people (age 65+), of which 40% live with a longstanding, limiting disease (1). An increase in physical disability and chronic disease among younger adults is expected to raise future demand for social care and disability benefits (2). Furthermore, the annual National Health Service (NHS) costs attributable to excess weight, a modifiable risk factor for multiple noncommunicable diseases, are projected to reach £9.7 billion by 2050 (3). The economic burden of disease prevention and treatment has been further amplified by the COVID-19 pandemic (4,5).

To mitigate the rising costs of health and social care, in 2011 the Department of Health released a new strategy that placed technological innovation at the forefront of policy making (6). The strategy’s objective was to harness the potential of technology to propel improvements in quality and efficiency (7). Areas where technological innovation proved to be particularly valuable include enabling early diagnosis (e.g., Healthy.io;(8)), empowering patients to gain control and choice in their treatment (e.g., Freestyle Libre; (9)), and facilitating remote monitoring for timely intervention (e.g., Video consultations; (10)).

Despite the advantages offered by implementing technological innovation in health and care, uptake in routine clinical practice has been generally poor. From 2009 to 2014, only 34% of technologies presented to the UK’s National Institute for Health and Care Excellence were recommended for NHS adoption (11). Adoption and diffusion of approved technologies is often slow, disrupted, and inequitable (12,13). For example, videoconferencing for remote consultations and monitoring is not new, but nation-wide attempts to its implementation prior to the COVID-19 pandemic have had limited success (14).

Successful implementation of innovative healthcare technology is dependent on its acceptability by health professionals and service users (15). Considering that the average lag time for technological innovation to be translated into routine practice is 17 years (16) and that this process requires significant investment, obtaining an early understanding of attitudes towards a technology in development can guide process evaluations and strategies to enhance and accelerate future acceptability of such technology. This will maximise the impact of initial investment and mitigate the risk of the technology becoming obsolete by the time it is adopted in routine care.

Research on digital health interventions (17,18) additionally shows that acceptability is highly variant between different groups of patients and the public, and it is heavily influenced by knowledge about the technology, perceived quality of the technology, and how the technology is endorsed. For example, uptake of tracking apps during COVID-19 was poor, despite the widespread use of mobile apps that normally monitor the user’s location, and this phenomenon was partially attributable to public distrust (19). Existing physical activity interventions utilising digital technology have been found to be much less effective in low compared to middle-high socioeconomic groups (20).

As researchers strive to create innovative technologies that enable targeted and timely interventions, the present study aimed to capture UK public opinions and attitudes to a range of novel healthcare technologies (NHTs). The specific objectives were: i) evaluate the public’s acceptability of these technologies in their future care; ii) discern the breadth of diversity in self-reported predispositions towards these technologies; and iii) seek initial insight into how these technologies could be introduced to patients in the future. Unlike previous research, we presented four different technologies in development, and asked respondents the same set of questions. By facilitating comparability between technologies, this study attempted to explore whether knowledge, attitudes and beliefs towards the introduction of new technology can be generalised to all NHTs or is contingent on the technology and its proposed application.

## Methods

### Study design and setting

A cross-sectional online survey.

### Population, sampling, and data collection

The survey was distributed online and managed by PureProfile, a professional social research company with over 140,000 registered individuals with a range of characteristics representative of the UK population geographically and demographically. Members of the public registered with PureProfile were eligible to participate if they were aged 18 years or above and lived in the UK. The survey was open for recruitment from the 30 April 2020 to 20 May 2020.

### Ethical approval

Ethical approval was obtained from the Faculty of Health Sciences Research Ethics Committee, University of Bristol (Ref: 94502).

### Survey development

Preliminary interviews were conducted by co-authors (HC, SG, BS, TP, SS, AV) with 12 healthcare professionals (HCPs) with diverse expertise, seniority, degree of patient contact. These interviews explored participants’ views and experiences of implementing NHTs in the UK national healthcare system, barriers and facilitators.

Finding from these interviews were used to draft the survey by all co-authors; an interdisciplinary team with expertise across the translational pathway of health technology development and implementation evaluation. Grounded on insight from the interviews, ‘novelty’ could be attributed to the actual technology underpinning a medical device or the health application of an existing technology. Four NHTs targeting well known and common chronic conditions were used in the survey as case studies. All were either being researched or commercial development phase at the time the survey was distributed. Table 1 displays technologies selected and differences/overlaps between them (See Supplementary Material 1 for details on the case studies).

**Table 1.** Characteristics of NHTs included in the survey.

|  | NHT 'type' | Function | Technology | Medical condition | Anticipated resistance to acceptance prior to the survey. |
| --- | --- | --- | --- | --- | --- |
| **Laser-based tool for early diagnosis** | Completely new | Early diagnosis | Nonlinear microscopy | Osteoarthritis (1 in 10 adults in UK;(21)) | No cure exists for this condition. |
| **Virtual reality to support self-management** | Novel application, existing technology. | Treatment | Virtual reality | Diabetes Mellitus/Obesity (7% of UK (22)/ 28% of adults in England(23)) | Public 'anxiety' around gaming. Ownership of data. Expectations of dizziness and tiredness from use. |
| **Continuous glucose monitoring using microwave signals (Heungjae, 2015)** | Completely new | Self-monitoring | Microwaves | Diabetes Mellitus (7% of UK population (22)) | Public 'anxiety' around microwave exposure<br>Myth of microwaves causing cancer<br>Relational memory linked to using domestic microwave ovens |
| **Mobile app for patient reported monitoring** | Novel application, existing technology. | Self-monitoring | Smartphone application | Rheumatoid arthritis (1% of UK population (24)) | Ownership of data<br>Privacy |

The structure of the survey comprised both multiple-choice and open questions across the following sections (See Supplementary Material 1):

- Familiarity with technology: Questions targeted overall technology use as well as use of technologies to manage own health.
- Presentation of case studies of the NHTs: First, survey items captured preliminary knowledge and predispositions towards the technology. Second, respondents were asked to indicate (on a Likert Scale) how likely they would agree to the use of the NHT after reading a) information about the NHT and its proposed application, b) the benefits, c) the risks. Three separate agreement scores were provided for each NHT, ranging between 1 (“Strongly Agree”) and 5 (“Strongly Disagree”). Finally, survey items captured respondent’s views on who and how the NHTs should be introduced to them.
- Experience with monitoring technology: Data were captured on use of this technology, opinions regarding access to information collected by the technology, and comfort in sharing sensitive information via the technology.
- Sociodemographic characteristics: Age, gender, ethnicity, index of maximal deprivation, education, general health.

Prior to launching the survey, an NIHR Bristol Biomedical Research Centre Patient and Public Involvement group piloted the survey and provided feedback. Two group sessions were run. The information was collated and incorporated in subsequent iterations of the survey.

The final draft of the survey was digitalised by the PureProfile team and piloted on 100 participants via their online platform. Modifications were made to the layout, format and logic of the questions prior to launch. The survey was distributed randomly by PureProfile, but quotes were applied for age and gender to ensure responders were a nationally representative sample. Respondents were provided with £2.10 payment by PureProfile for the time taken to complete the survey. Anonymised survey data were provided to the study team for analysis and the final sample size included the pilot responses.

### Statistical analysis

Summary statistics show participant responses to survey questions. For case study responses, a single agreement score was calculated by averaging the three agreement scores provided. A lower agreement score was indicative of a higher degree of agreement to accepting the NHT in their future care. All three scores were required to calculate the average, otherwise the agreement score was left missing for the participant. Index of Multiple Deprivation (IMD) Quintile was determined from responders’ postcodes provided in the survey (See Supplementary Material 2 for details).

Two open-ended questions were included in the survey to explore respondents’ reluctance to accept the NHTs presented in the case studies and views on how health professionals should introduce the presented NHTs to patients in the future. These open-ended free text questions were analysed using content analysis. The first 50 responses to the two questions were coded independently by the research team (SS, AV, HC, TP for question 1; SS, SeS, HC, GL for question 2). A coding framework was developed, after which researchers coded the remaining responses. SS further coded 10% of all responses to check for consistency. Frequency that codes emerged in responses were calculated.

Data were provided from PureProfile in IBM SPSS Statistics 25 and all descriptive analyses were undertaken in this package.

## Results

Preliminary interviews with HCPs revealed that the survey would have to provide participants with a definition of ‘Novel Healthcare Technology’ and that the technology should be presented alongside its proposed application (i.e., the medical condition). HCPs highlighted that at present there is a lack of standard procedure through which patients are introduced to NHTs and there is a need to identify and adequately train the key people involved in this process. This information was used to shape the survey and incorporate additional questions around respondents’ views on how NHTs should be introduced to future patients.

### Sample

A total of 1,450 people responded to the survey. Median time taken to complete the survey was 15.60 minutes (range: 4.77 mins to 1958.5 minutes). Respondents were based across the United Kingdom, though most were from London (13.2%) and the South East England (13.9%).

Mean age of participants was 46.4 years (SD: 17.13) and 50.6% were female. The majority of respondents were British White (80.1%) and the overall distribution of the sample paralleled population data on ethnicity collected by the Office of National Statistics (ONS, 2011). Most respondents were in active paid work (56.4%), 32.1% held a University degree, and IMD score was evenly spread across quantiles. Respondents mainly rated their general health as ‘Good’ (57.2%), and only 3.5% indicated ‘Bad’ or ‘Very Bad’. 63.9% of participants indicated they had no health condition, with most participants who reported a health condition classifying it as ‘long-term illness, disease or condition’ (14.4%). (Supplementary Material 2 for details).

### Knowledge and use of NHTs

Overall, a large majority of participants indicated they were frequent users of technology in their everyday life (82.6%), though only 9.4% reported that novel technologies were being used for management of their health. Of respondents who had previously used NHTs (n = 225), most made use of these technologies every few months (25.3%) or once every few years (27.1%).

Regarding the NHT case studies included in the survey, Table 2 displays participants’ previous understanding of how the actual technology works. Self-reported knowledge on laser-based and microwave-based technology primarily ranged between ‘some understanding’ to ‘never heard of them’, while responses were more diverse for mobile app and virtual reality technology. Yet, awareness of the use of these technologies in healthcare was highest for mobile apps (66%), followed by laser-based technology (63.8%), microwave technology (28%) and lastly virtual reality (18.3%).

**Table 2:** Previous awareness or use of the NHTs included in the survey case studies

| Item | Category | Laser-based tool for early | Virtual reality to support | Continuous glucose | Mobile app for patient |
| --- | --- | --- | --- | --- | --- |

|  |  | diagnosis |  | self-management |  | monitoring using microwave signals |  | reported monitoring |  |
| --- | --- | --- | --- | --- | --- | --- | --- | --- | --- |
|  |  | n | % | n | % | n | % | n | % |
| How much do you understand? | Never heard of them | 437 | 30.1 | 196 | 13.5 | 609 | 42.0 | 317 | 21.9 |
|  | Have heard but don't understand | 411 | 28.3 | 246 | 17.0 | 322 | 22.2 | 157 | 10.8 |
|  | Have some understanding | 499 | 34.4 | 601 | 41.4 | 343 | 23.7 | 395 | 27.2 |
|  | Understand quite well | 85 | 5.9 | 306 | 21.1 | 138 | 9.5 | 333 | 23.0 |
|  | Understand and could explain to others. | 18 | 1.2 | 101 | 7.0 | 38 | 2.6 | 248 | 17.1 |
| Are you aware of use in healthcare? | Yes | 925 | 63.8 | 266 | 18.3 | 406 | 28.0 | 957 | 66.0 |
|  | No | 525 | 36.2 | 1184 | 81.7 | 1044 | 72.0 | 493 | 34.0 |

### Acceptability of NHTs in future care

After reading information about the NHTs and the proposed health application, including associated benefits and risks, median acceptance score provided by respondents was 2.0 for virtual reality to support self-management (Interquartile Range (IQR): 1.3-3.0) and continuous glucose monitoring using microwave signals (IQR: 1.3-2.3), and 1.7 for laser-based technology for early diagnosis (IQR: 1.3-2.0) and mobile app for patient reported monitoring (IQR: 1.0-2.0). Agreement scores ranged between 1 “Strongly Agree” to 5 “Strongly disagree”. Table 3 for details.

**Table 3.** Reasons (frequency of mention) for not accepting the NHTs as part of their care. Bold denotes most frequent reason provided.

| Item | Category | Laser-based tool for early diagnosis |  | Virtual reality to support self-management |  | Continuous glucose monitoring using microwave signals |  | Mobile app for patient reported monitoring |  |
| --- | --- | --- | --- | --- | --- | --- | --- | --- | --- |
|  |  | n | % | n | % | n | % | n | % |
| If you would still not use the technology, why not? | Don't know | 62 | 18% | 58 | 10% | 87 | 15% | 70 | 27% |
|  | Provided a reason: |  |  |  |  |  |  |  |  |
|  | More info | 61 | 18% | **106** | **18%** | 60 | 11% | 56 | 21% |
|  | Safety | 46 | 13% | 67 | 12% | **138** | **25%** | 1 | 0% |
|  | Evidence | 28 | 8% | 67 | 12% | 67 | 12% | 1 | 0% |
|  | Risks | **71** | **21%** | 23 | 4% | 52 | 9% | 6 | 2% |
|  | Long term | 11 | 3% | 2 | 0% | 88 | 16% | 0 | 0% |
|  | Side Effects | 18 | 5% | 47 | 8% | 29 | 5% | 1 | 0% |
|  | Other | 7 | 2% | 60 | 10% | 2 | 0% | 11 | 4% |
|  | Test Technology | 0 | 0% | 45 | 8% | 20 | 4% | 1 | 0% |
|  | Privacy | 0 | 0% | 1 | 0% | 1 | 0% | **60** | **23%** |
|  | Trust | 3 | 1% | 27 | 5% | 5 | 1% | 10 | 4% |
|  | Cost | 12 | 3% | 25 | 4% | 3 | 1% | 3 | 1% |
|  | HCP info | 15 | 4% | 13 | 2% | 9 | 2% | 4 | 2% |
|  | Individual risk | 9 | 3% | 13 | 2% | 1 | 0% | 6 | 2% |
|  | Access | 0 | 0% | 4 | 1% | 0 | 0% | 24 | 9% |
|  | Adjunct | 0 | 0% | 17 | 3% | 0 | 0% | 3 | 1% |
|  | Intrusive | 0 | 0% | 0 | 0% | 0 | 0% | 6 | 2% |

### Acceptability according to individual characteristics

Using cross tabulation, several patterns were detected in acceptance according to respondent characteristics. Increasing age was linked with an increased willingness to adopt laser-based technology for early diagnosis and a mobile app for patient reported monitoring (Figure 1). Age had no effect on acceptance scores for the other NHTs. Presence of a self-reported condition followed a similar pattern, having a positive effect on acceptance in the case studies depicting laser-based technology and mobile apps, but not the other NHTs (Figure 2). A gender difference was only found in acceptance of mobile apps for patient reported monitoring, being slightly elevated in females (median: 1.3; IQR: 1.0-2.0) compared to males (median: 1.7; IQR: 1.0-2.0). Agreement ratings towards the mobile apps were also closer to “Strongly Agree” for those aware of the use of these apps in healthcare (median: 1.3; IQR: 1.0-2.0) compared to those not aware (median: 2.0; IQR: 1.0-2.3). These patterns were not observed for the other NHTs (Supplementary Material 3).

**Figure 1:**
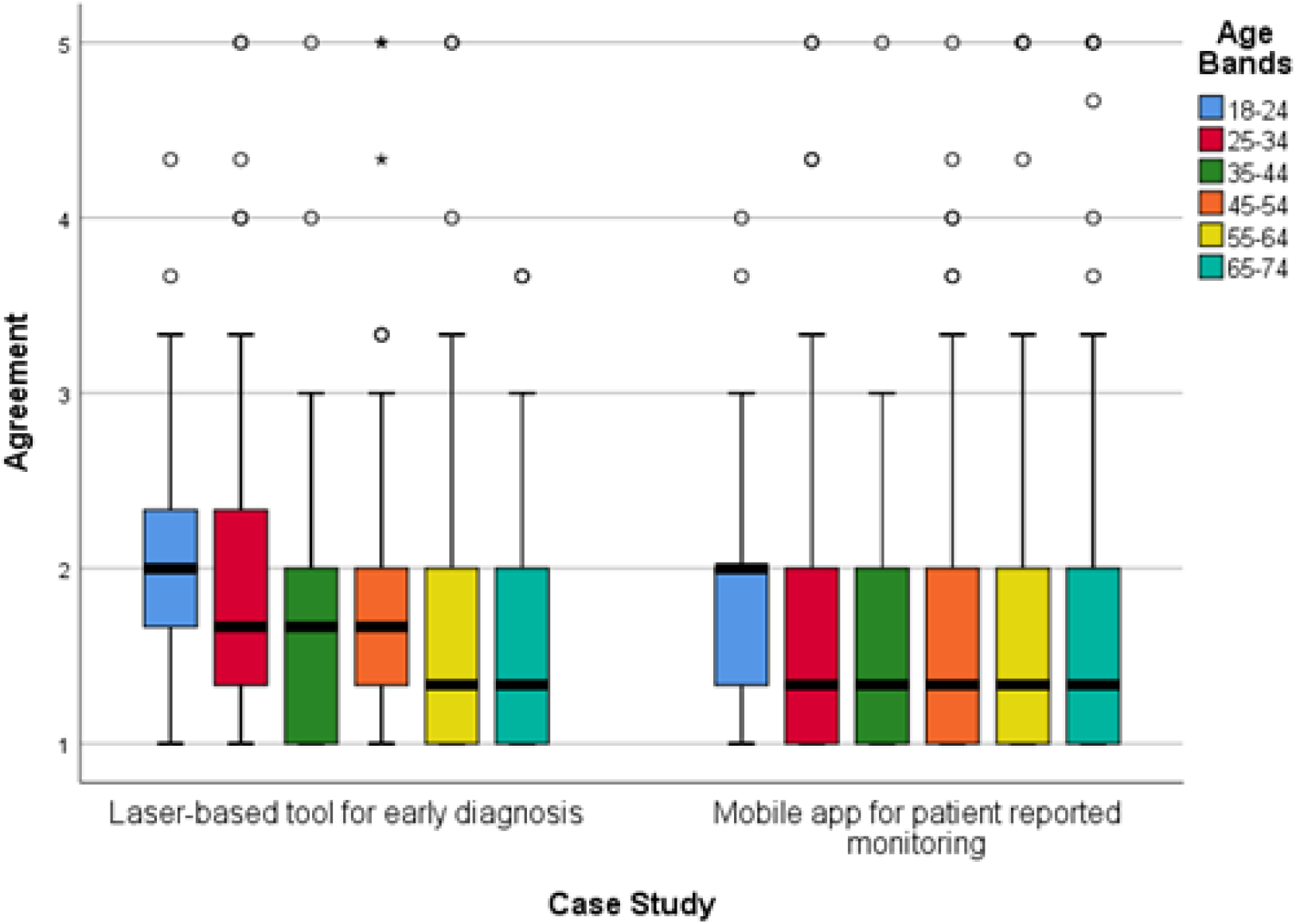
Degree of agreement to accept the laser-based technology and mobile apps in future care according to respondents’ age. Agreement score ranges between 1 “Strongly Agree” to 5 “Strongly disagree”. Circles indicate mild outliers (between 1.5 and 3 interquartile ranges away from the 75^th^ percentile) and stars indicate extreme outliers (greater than 3 interquartile ranges away from the 75^th^ percentile). Laser-based tool for early diagnosis n = 1139. Mobile app for patient reported monitoring n = 1093.

**Figure 2:**
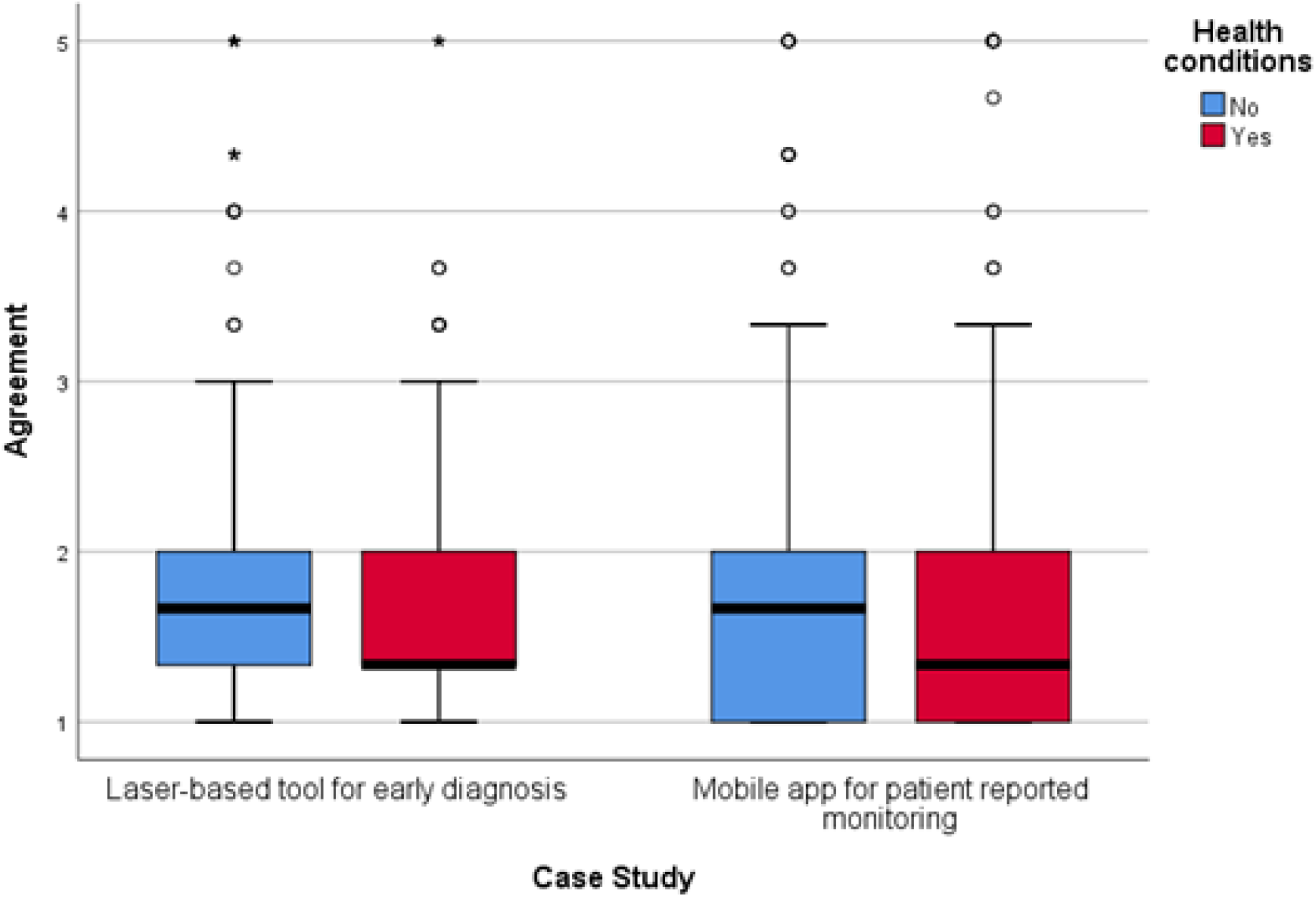
Degree of agreement to accept laser-based technology and mobile apps in future care, split by respondents who reported having a medical condition and those who didn’t. Agreement score ranges between 1 “Strongly Agree” to 5 “Strongly disagree”. Circles indicate mild outliers (between 1.5 and 3 interquartile ranges away from the 75^th^ percentile) and stars indicate extreme outliers (greater than 3 interquartile ranges away from the 75^th^ percentile). Laser-based tool for early diagnosis n = 1139. Mobile app for patient reported monitoring n = 1093.

As shown in Figure 3, being a frequent user of technology in everyday life impacted acceptability of all NHTs except for continuous glucose monitoring using microwave signals. Respondents’ self-reported understanding on how the various NHTs work was also linked to acceptability. Across all NHTs, those responders with a self-reported higher baseline understanding (i.e., “Understand and could explain to others”) about how the technology worked were more likely to “Strongly Agree” to their use when compared to other responders. (Figure 4).

**Figure 3:**
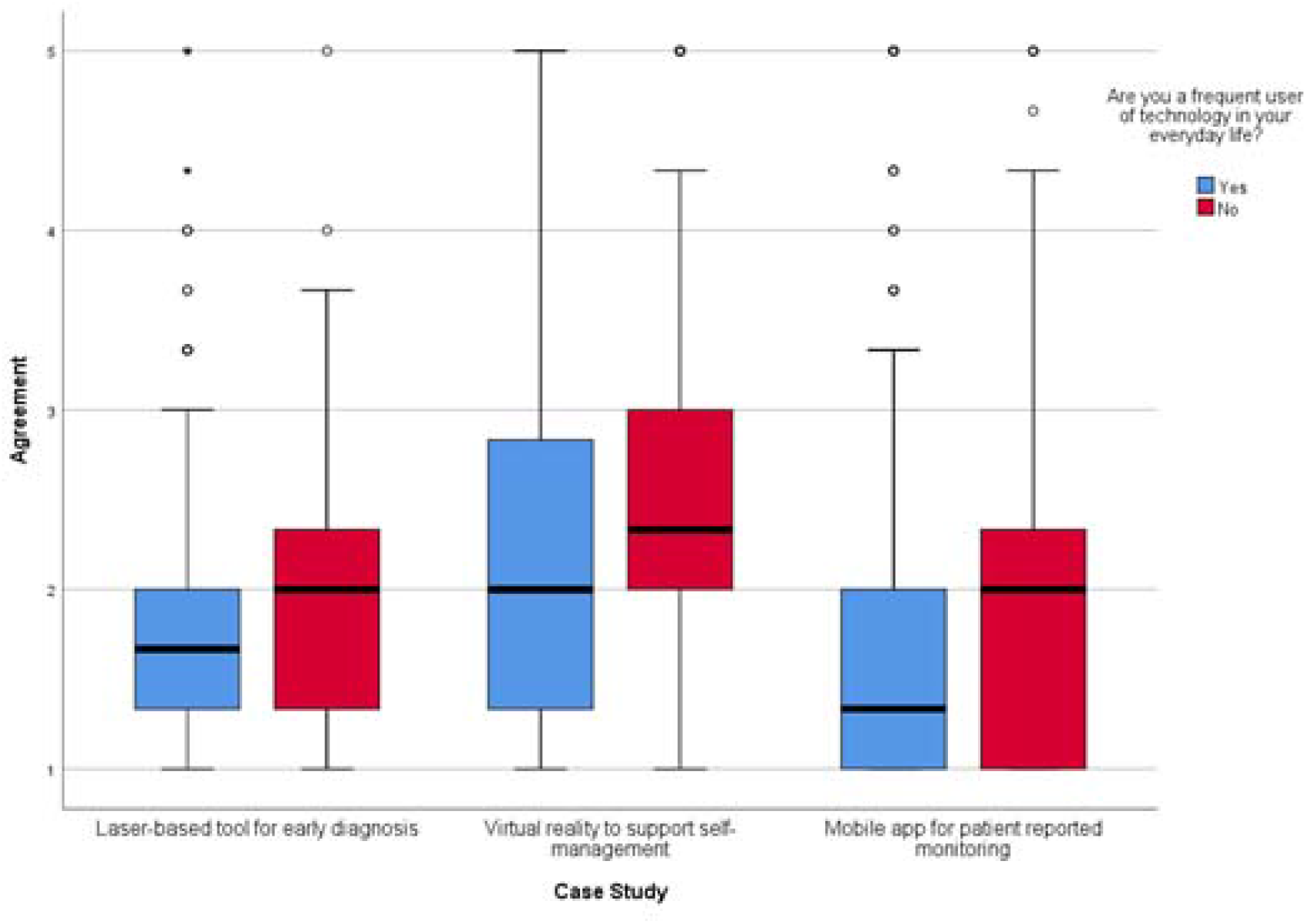
Agreement to accepting laser-based technology, virtual reality and mobile apps in future care, split by respondents who reported being frequent users of technology in everyday life and those who were not frequent users. Agreement score ranges between 1 “Strongly Agree” to 5 “Strongly disagree”. Circles indicate mild outliers (between 1.5 and 3 interquartile ranges away from the 75^th^ percentile) and stars indicate extreme outliers (greater than 3 interquartile ranges away from the 75^th^ percentile). Laser-based tool for early diagnosis n = 1139. Virtual reality to support self-management n = 1089. Mobile app for patient reported monitoring n = 1093.

**Figure 4:**
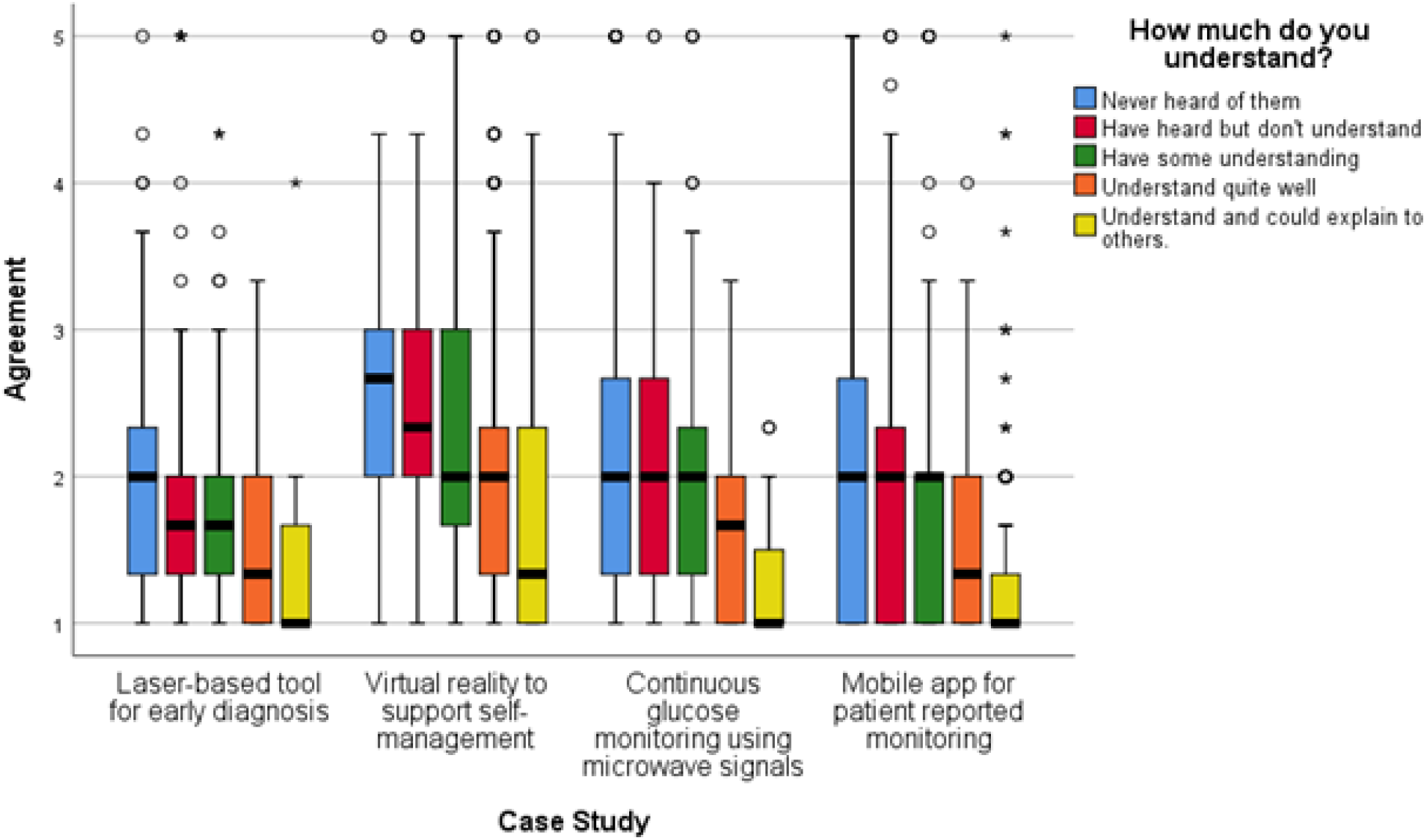
Agreement to accepting NHTs in future care according to respondents’ perceived understanding of how the technology works. Agreement score ranges between 1 “Strongly Agree” to 5 “Strongly disagree”. Circles indicate mild outliers (between 1.5 and 3 interquartile ranges away from the 75^th^ percentile) and stars indicate extreme outliers (greater than 3 interquartile ranges away from the 75^th^ percentile). Laser-based tool for early diagnosis n = 1139, Virtual reality to support self-management n = 1089, Continuous glucose monitoring using microwave signals n = 1119, Mobile app for patient reported monitoring n = 1093.

No patterns were observed in acceptability in relation to ethnicity, IMD quintile, education, self-reported general health, and self-reported experience with NHTs for health management (Supplementary Material 3).

### Reluctance to accept

When querying reluctance to accept the NHTs after providing information, risks and benefits of each NHT and its proposed application, differences were found between case studies in the reasons for expressing hesitation (Table 3). Between 10% (virtual reality) and 27% (mobile app) of respondents did not know why they were unwilling to accept the technology after reading the information provided. Overall, participants wanted more information about the NHTs, expressed concerns regarding the risks and safety of using the NHTs, and wanted more scientific and clinical evidence before they would be willing to accept them as part of their care.

For the laser-based technology, the most common concern was the risk attached to undergoing surgery. Respondents questioned whether *“benefits would outweigh the risks”* (P622), the *“risk associated when not used properly”* (P1400), and *“risk of localised damage”* (P1435). Respondents therefore also requested more detailed information on the procedure, effectiveness, and risks involved.

The most frequent concern expressed by participants regarding the use of virtual reality to support self-management was the need of additional information about the technology and how it worked. Some respondents struggled to see the benefits of using the NHT, or the added value to more traditional approaches to diabetes management, *“I don’t understand what is involved, the time commitment, why it is better than me just eating less and exercising more or the evidence of efficacy”* (P764). Respondents expressed wanting more evidence regarding its effectiveness and implementation. Concerns were also raised about safety, with participants being weary of the possibility of feeling “nauseous”, “sick”, “dizziness” and “disorientated” when using virtual reality. Consequently, the opportunity to test this technology played a more important role in the decision-making around acceptability: “*I would base my final decision on the results of a demonstration and further information”* (P308)

Similarly, reluctance to accept continuous glucose monitoring using microwave signals was primarily attributable to perceived safety and the long-term effects. Participants expressed particular concern about the impact of microwaves on health. However, respondents also demonstrated that evidence would be an important factor in their decision-making: *“I would like more research done first to show me it’s safe”* (P518)

In the use of mobile apps for patient reported monitoring, concerns about privacy were the most frequently reported reason for non-acceptance, which were not present with the other NHTs. Respondents expressed *“reservations about data security”* (P207), *“privacy concerns over who would have access to the information”* (P881) and *“using of personal information without acknowledgement”* (P1386). Again, respondents indicated that more information would need to be provided before they would accept this NHT (%): *“I would use it if someone took time to explain it fully to me”* (P291)

### Approaches to introduce NHTs to patients

Across all NHTs, most participants (range 53.6% to 65%) preferred receiving information about the NHT verbally, with respondents wanting the information to come from a healthcare specalist or a general practitioner (see Table 4). The second most preferred form of receiving information was via a webpage across all NHTs except virtual reality, where providing information via a visual demonstration was selected to a similar degree to having a verbal conversation about the NHT.

**Table 4.**
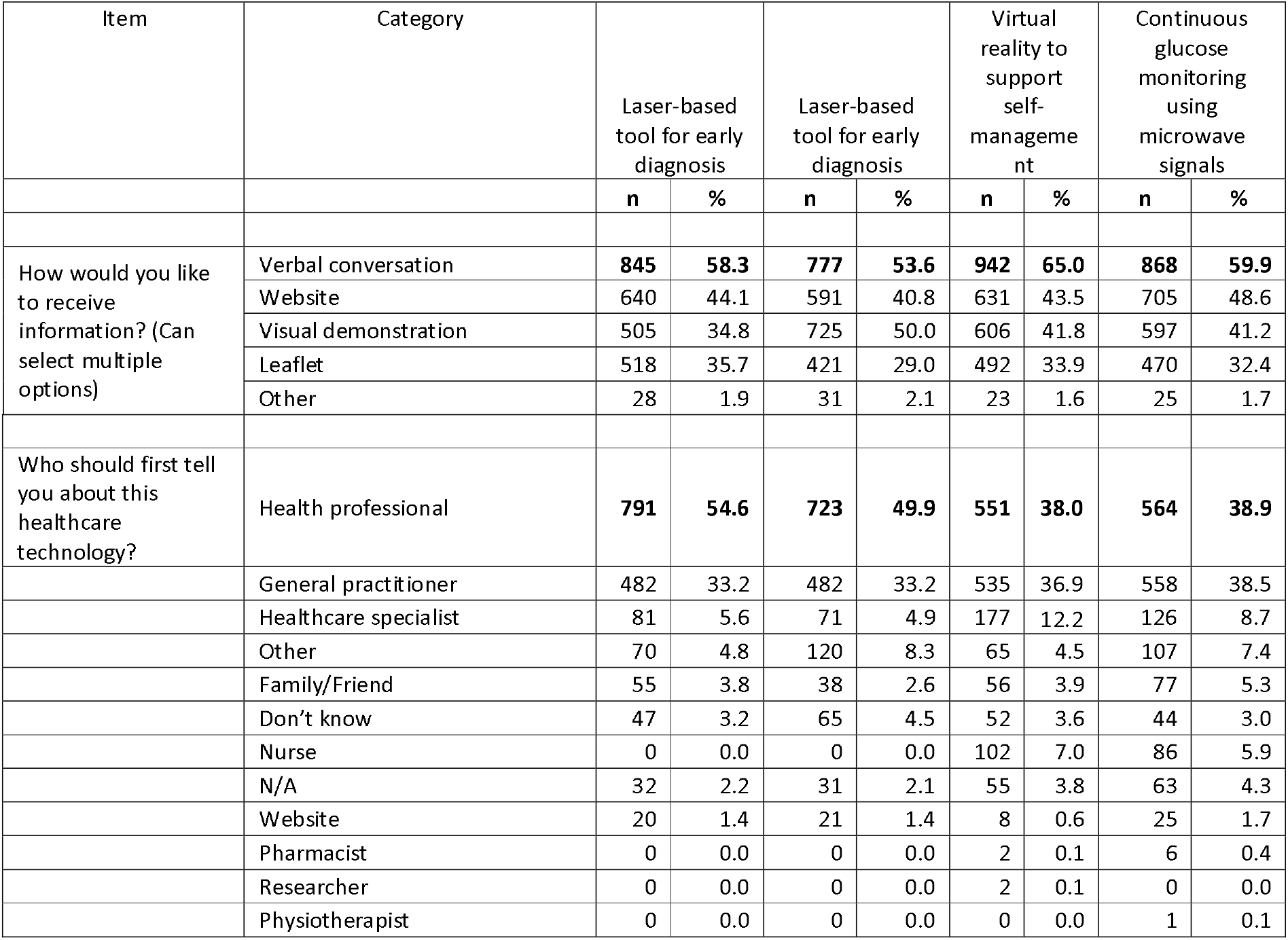
Method and ideal person to introduce a NHT to future patients.

## Discussion

Acceptance of an innovative technology is contingent on the attitudes of those who will be using the technology (25,26). This survey accrued the public’s attitudes towards NHTs in development; a laser-based tool for early diagnosis, a virtual reality tool to support self-management, a non-invasive continuous glucose monitor using microwave signals, a mobile app for patient reported monitoring. Responses showed that self-reported knowledge about NHTs is diverse, with baseline awareness being highest for mobile apps, followed by laser-based technology, microwave technology and lastly virtual reality. Acceptability of future use varied across the four NHTs, with the highest median acceptability scores achieved for the mobile app and the laser-based technology. Respondents’ self-reported understanding of how the various NHTs worked increased acceptability across all technologies, and frequent users of novel technologies also displayed greater acceptability in all NHTs except for the microwave-based technology. Hesitation about adopting the NHTs was expressed by some participants. Key concerns related to the risks associated with the NHTs and safety. For those respondents who remained hesitant about adopting the NHTs, more information was needed to help inform their decisions, with scientific and clinical evidence the most commonly requested evidence.

Aligned with the literature (17), respondents who reported greater understanding of how a particular NHT works also provided higher acceptance scores to that NHT. Similarly, respondents were more familiar with the healthcare application of laser-based technology and mobile apps, which were also the NHTs that received the highest acceptance scores. These findings parallel research conducted with HCPs showing that perceived lack of technical competency and unfamiliarity generates reluctance to try novel digital solutions (13), and providing ‘how-to’ knowledge at early stages of innovation should be reinforced to enable implementation success (27). The application of knowledge transfer strategies as done at an organisational level (e.g., 27) is likely to be valuable to stimulate a positive attitude towards NHTs.

In contrast to previous work on digital health (e.g., 17) no trends were found in acceptance scores when considering respondents’ ethnicity, socioeconomic status or level of education. Acceptability scores were higher for female respondents and those who reported frequent use of technology only in some case studies. Further, there was greater acceptability for the mobile app and laser-based technology among the older respondents, who are typically associated with adversity to technology (29). These findings suggest that we cannot make assumptions about prospective acceptability of a premature NHT based on evidence of similar, existing solutions. The discrepancy between results obtained in this study and previous research may reflect the importance of perceived usefulness and ‘felt need’ on older adults’ valuation of NHTs (29–31); arthritis is generally an age-related disease for which there is no cure but the proposed NHTs can help slow down its progression through timely action. Though older adults may be more adverse to technological solutions, the elevated perceived threat of arthritis may make them more likely to accept NHTs that directly target this condition.

When endorsing a NHT to a patient, information about the NHT should match the preconceptions and knowledge the individual might have of the technology. Safety was an important concern regarding both the laser-based and microwave-based technologies. However, while for laser-based technology participants wanted more information on the surgical procedure and its effectiveness, for the latter there was a call for more evidence on the wider effects of microwaves on human health. This may reflect the high prevalence of incorrect beliefs found on the causal impact of non-ionizing electromagnetic frequencies on cancer risk (32); misconceptions about microwave signals could have impacted acceptability of its use for glucose monitoring as found in this study. The survey also showed that privacy concerns contribute to reluctance on accepting a novel intervention using a mobile app, but not one relying on virtual reality, which is also vulnerable to disclosure of personal data (33). The literature on these technologies applied to other health conditions shows a similar trend (see 33,34). Information provided about an NHT needs to be tailored to address existing preconceptions, rather than a brief outline as provided in this study.

A considerable strength of this study is the use of a large-scale survey to obtain insight on attitudes towards innovative technological solutions that are still at early stages of development. Both the people designing and developing these NHTs (including authors of this manuscript) and implementation scientists can use results to prospectively mitigate obstacles to acceptance and adoption. For example, respondents who struggled to understand how the virtual reality works also showed greater reluctance to accept the technology when presented with its possible application to support diabetes self-management. Results from this survey suggest that enabling target beneficiaries to try the technology before beginning to use the intervention fully could increase acceptability. The value of inserting a ‘trial phase’ when implementing this technology aligns with evidence that older adults change their attitudes towards virtual reality after using it (34). Further, the survey evaluated four very different NHTs that target different healthcare challenges, enabling comparability between the technological solutions and highlighting the importance of NHT decision-making to be conducted on a case-by-case basis (i.e., each technology and its application).

There are, however, certain limitations that need to be considered. The technological solutions presented in this study varied across different factors (e.g., medical application, function). This study was based on an online survey, which means that the attitudes of people who do not normally use the internet were not captured. The majority of respondents did not report a diagnosis of the medical conditions presented, which is likely to influence how likely individuals would accept a technology if they were experiencing the adverse symptoms of a condition. The survey also explored attitudes and prospective acceptability of NHTs. Though acceptance is closely linked to behavioural intention of using technology (26), it cannot account for experienced usability (25,36). Further research is needed to evaluate how the outcomes from this survey reflect future adoption of the NHTs when implemented.

### Practical Implications

This study emphasises the complex task stakeholders face in the development and implementation of NHTs, where it is impossible to predict attitudes to future NHTs on the basis of existing solutions. Insight into the public’s attitudes towards proposed NHTs can guide product development and early identification of implementation strategies (12,37). Understanding public attitudes towards proposed technological solutions can assist funders and regulatory agencies in their decision-making, and the preparation of blueprints for introducing NHTs in routine practice that can adapt to the type of technology, its application and the users of the technology. It can also assist researchers ensure that innovation is inclusive, and that sustainable support is in place to assist future beneficiaries access the healthcare solution (13,38). Additionally, the relevance of an individual’s understanding of the NHTs and preference for direct conversations with health professionals strongly supports the adoption of shared decision-making practices (39) to increase acceptance of NHTs in health and social care.

## Conclusion

This study examined UK public attitudes and opinions towards NHTs being used to address common chronic conditions. Responses suggest that the UK public is generally open to technological innovation in their future care but acceptability of a specific NHT is likely to vary. When considering factors that might explain such variation (e.g., age, gender, familiarity with technology), this study highlighted the need to avoid generalisations across technology; the relevance of individual/group characteristics on acceptability differed across case studies. Similarly, reluctance to accept the NHTs was driven by differing rationales, which might be linked to the individual’s understanding of the technology, or the perceived balance of benefits versus costs of using the NHT as a targeted solution for a specific clinical need. By adopting a research framework that encompasses several NHTs in development, this study evidences the complexity underpinning acceptability research in the field of healthcare technology innovation. A comprehensive evaluation of how future beneficiaries might respond to a technological solution as it is being conceptualised, has potential to streamline the development and implementation processes and maximise its health and societal impact.

## Data Availability

All data produced in the present study are available upon reasonable request to the authors

## List of Abbreviations

NHT: Novel healthcare technology
HCP: Healthcare professional
IMD: Index of Multiple Deprivation

## Declarations

### Ethics approval and consent to participate

Ethical approval was obtained from the Faculty of Health Sciences Research Ethics Committee, University of Bristol (Ref: 94502). Consent to participate was provided by respondents by completing the survey.

### Consent for publication

Not applicable

### Availability of data and materials

The datasets used and analysed during the current study are available from the corresponding author on reasonable request.

### Authors’ contributions

SS, TP, AV, HC, BS, JZ, SC, SG and GL conceptualised the research question, developed the study design, designed the survey and obtained funding to support the research activities. SS, TP, AV, BS and SG interviewed the semi-structured interviews that informed the survey, and SS ran the PPI activity. GL co-ordinated the acquisition of the data, while SS, TP, AV, HC, JZ and SS analysed the data. All researchers were involved in the interpretation of data. SS drafted the manuscript, and the rest of the team made substantial contributions to iterations of the manuscript. All authors approve the submitted version and have agreed both to be personally accountable for their own contributions and ensuring that questions related to the accuracy or integrity of any part of the work are appropriately investigated, resolved and the resolution documented in the literature.

## Acknowledgements

We thank the NIHR Bristol BRC Diabetes Patient and Public Involvement Group for their contribution to the design of the survey.

