## Supplementary Material 1 for "Public attitudes towards the use of novel technologies in their future healthcare: A UK survey"

**Supplementary Material 1 – Survey and case studies presented on the novel healthcare technologies selected**

### GW4 PATH: Perceptions and Attitudes of Technologies in Healthcare

This survey has been designed by a group of researchers from the universities of Bristol, Bath, Exeter, Cardiff and Worcester. They are looking at people’s views on the use of novel technologies in healthcare.

Completion of this questionnaire is anonymous. You are free to withdraw at any point by exiting the survey. However, any data collected up to this point will be retained. All data will be stored securely and is covered by the Data Protection Act 2018. By completing this survey, you are consenting to the researchers using your anonymous responses in their research.

--	--	--	--	--	--	--	--	--	--	--	--	--

#### Novel technologies in the NHS are things like Apps, programmes, medical devices or diagnostic tests that are not yet part of standard care across the NHS. Specific examples include robotic surgery, artificial intelligence, nurse robots.

Novel technologies differ from routine technologies, which are tools or procedures that are widely used by healthcare professionals. Example of routine technologies are finger-prick tests for diabetes, x-rays to examine bone fractures, pacemakers to monitor heart rhythm.

#### Section 1

1. Are you a frequent user of technology in your everyday life?

- Yes
- No

2. Have novel technologies been used in the management of your health?

- Yes
- No
- Unknown

3. How often do you have contact with these new technologies *for the management of your health*?

- Never
- Once every few years
- Once a year
- Every few months
- Every month
- Every week
- Every day

4. What novel technology do you use the most?

____________________________________________________________________________________________________________________________________________________________________

5. Think about the novel technology in healthcare that you use the most:
   5.1. Who first gave you information about this novel technology?

- Personal choice
- Family/friends
- GP/doctor
- Nurse
- Other ________________________
  5.2. Before I started using this novel technology I believed it would be easy to use
- Strongly agree
- Agree
- Neutral
- Disagree
- Strongly disagree

5.3 What do you think about this novel technology now that you use it?

______________________________________________________________________________________________________________________________________________________________________________________________________________________________________________________

5.4 What made you decide to use this novel technology? [select all that apply]

- It would be more useful/better than routine care.
- My doctors/nurses gave me all the information that I needed.
- Easy to access and use.
- It would improve my physical appearance.
- It looked nicer.
- I was given money to be part of a study about the technology.
- I trust everything my doctors recommend.
- I was part of a study that could help other patients.
- I could avoid long waiting list.
- Research shows it works.
- My doctor told me that the clinical team has used the technology before.
- Not sure
- Other__________________________________________
  1.5 On reflection, if you wanted to change anything about how this novel technology was introduced to you, what would it be?

__________________________________________________________________________________

__________________________________________________________________________________

__________________________________________________________________________________

#### Section 2 – DIAGNOSIS

**You will now read a case study of a new technology being developed for the early diagnosis of a medical condition. Please imagine you are one of the patients.**

Case study 1: Early detection of osteoarthritis – A laser-based tool for diagnosis

- You suffer a knee injury that has a 50/50 chance of causing an accelerated form of arthritis.
- Current medical imaging technology can’t detect the presence of arthritis until it has severly damaged your knee.
- Your doctor tells you about a new imaging technology that uses a low power laser to look for microscopic signs of early stage arthritis.

a) How much do you understand about laser-based technologies work? [tick one]

- Never heard of them
- Have heard but don’t understand
- Have some understanding
- Understand quite well
- Understand and could explain to others.

b) Are you aware that laser-based technologies are used in healthcare?

- Yes
- No

c) How do you feel about laser-based technologies being used in healthcare?

________________________________________________________________________________________________________________________________________________________________________________________________________________________________________________________________________________________________________________________________________

**Information 1: Why?**

- Arthritis is a disease of the joints that causes chronic pain and affects your ability to do everyday things like walking, writing and driving.
- Early detection of arthritis is key for minimising the damage it does to the joint. However, the best techniques used by doctors to detect arthritis are only sensitive to the condition at an advanced stage.
- The technology introduced here has the potential to detect arthritis at the earliest possible stage, often decades before it can be detected today.

d) After reading this information about the case study, I would accept the use of laser-based tool for early diagnosis of early stage arthritis.

- Strongly agree
- Agree
- Unsure
- Disagree
- Strongly disagree

**Information 2: Benefits**

- Early detection of arthritis will significantly reduce the chances of you needing joint replacement surgery.
- The laser-based imaging tool can be used during routine key-hole operations, meaning no additional visits to the hospital are required.

e) After reading this information about the case study, I would accept the use of laser-based tool for early diagnosis of early stage arthritis.

- Strongly agree
- Agree
- Unsure
- Disagree
- Strongly disagree

**Information 3: Risks**

- This technology uses an infrared laser beam to capture microscopic images of your joint. Shining too much laser light on biological tissues can cause damage (a bit like too much sun is not good for your skin).
- There is a small risk that some localised damage to your joint tissues may occur.

f) After reading this information about the case study, I would accept the use of laser-based tool for early diagnosis of early stage arthritis.

- Strongly agree
- Agree
- Unsure
- Disagree
- Strongly disagree

g) **If you are still unsure about whether you would accept the use of this technology, what other information would help you make a decision?**

- Research studies in humans
- Successful case studies of other patients
- Demonstration of how it works
- Other. Please state: __________________________________________________________________________________________________________________________________________

h) If you would still not use the technology, why not?

____________________________________________________________________________________________________________________________________________________________________

i) How would you like to receive the above information? [tick as many as apply]

- Leaflet
- Visual demonstration
- Verbal conversation with healthcare professional
- Website
- Other: ________________________

j) Who should first tell you about the possibility of using laser-based technology to screen for osteoarthritis?

_____________________________

k) Currently there are no approved treatments that will completely stop the progression of osteoarthritis. Would you still be interested in getting early diagnosis via this novel, laser-based healthcare technology?

- Yes
- No

#### Section 3 – TREATMENT

**You will now read a case study of a new technology being developed to support treatment of chronic medical conditions. Please imagine you are one of the patients.**

Case study 2: Virtual reality for type 2 diabetes.

You have been told by your doctor that you have type 2 diabetes and recommends that you lose some weight. Alongside general guidelines about how to lose weight through increasing physical activity and nutrition, the doctor suggests they are trialling the use of virtual reality as a training tool to support weight loss.

a) How much do you understand about how virtual reality works? [tick one]

- Never heard of them
- Have heard but don’t understand
- Have some understanding
- Understand quite well
- Understand and could explain to others.

b) Are you aware that virtual reality is used in healthcare?

- Yes
- No

c) How do you feel about virtual reality being used in healthcare?

________________________________________________________________________________________________________________________________________________________________________________________________________________________________________________________________________________________________________________________________________

**Information 1: Why?**

- Weight loss from lifestyle change can help people achieve diabetes remission.
- Weight loss interventions are time-limited and expensive. Most patients regain the weight lost.
- Virtual reality can help people acquire skills to help them maintain dietary change and exercise regularly in the longer term.

d) After reading this information about the case study, I would accept virtual reality as a tool to help treatment.

- Strongly agree
- Agree
- Unsure
- Disagree
- Strongly disagree

**Information 2: Benefits**

- Patients receive more regular support for longer.
- The support provided can be adapted to the needs of each individual patient.
- The tool can train people to use specific techniques that NHS diabetes care teams do not have the resources to provide.
- Guidance/training is provided by showing real-life examples. This makes it easier to apply what is learnt to everyday life.

e) After reading this information about the case study, I would accept virtual reality as a tool to help treatment.

- Strongly agree
- Agree
- Unsure
- Disagree
- Strongly disagree

**Information 3: Risks**

- Some people experience nausea and dizziness when they first try virtual reality.
- Extensive use can cause eye strain.
- Can trigger seizures in people with epilepsy.

f) After reading this information about the case study, I would accept virtual reality as a tool to help treatment.

- Strongly agree
- Agree
- Unsure
- Disagree
- Strongly disagree

g) **If you are still unsure about whether you would accept the use of this technology, what other information would help you make a decision?**

- Research studies in humans
- Successful case studies of other patients
- Demonstration of how it works
- Other. Please state: __________________________________________________________________________________________________________________________________________

h) If you would still not use the technology, why not?

____________________________________________________________________________________________________________________________________________________________________

i) How would you like to receive the above information? [tick as many as apply]

- Leaflet
- Visual demonstration
- Verbal conversation with healthcare professional
- Website
- Other: ________________________

j) Who should first tell you about the use of virtual reality to help treat a medical condition?

__________________________________________________________________________________

#### Section 4 – Self monitoring of health using technology

The NHS is encouraging the use of mobile phone applications (Apps) and websites to help patients monitor their own health. Examples of self-monitoring technologies include things like Fitbit, Digital Health Passport App.

1. Are you actively monitoring your own health?

- Yes
- No
  1.1. If yes, how are you tracking it? [tick all that apply]
- Mobile application
- Website
- Wearable devices
- Others __________________________________
  1.2. How frequently do you use the technology to monitor your progress?
- Never
- Once every few years
- Once a year
- Every few months
- Every month
- Every week
- Every day

2. When using these technologies, patients are normally asked to enter general information about themselves (e.g. age) and more specific information relevant to their condition (e.g. eating habits and medical symptoms). Who do you think should be allowed access to this information? [tick all that apply]

- GP
- Specialist doctor
- Family
- NHS database
- Technology developers (if the data is anonymised)
- Researchers (if the data is anonymised)
- Other _______________________________

3. How comfortable are you in inserting sensitive information (e.g. weight, menstrual cycle) in the technology? Place a cross on the horizontal line.

Not at all ----------------------------------------------------------------------------------------------------------Completely

**You will now read case studies of new technologies being developed to monitor specific medical conditions. Please imagine you are one of the patients.**

Case study 3: Continuous glucose monitor for diabetes – Monitoring with microwave signals

You have diabetes, and your doctor gives you guidelines on how you need to monitor your blood sugar level every day. This can be done by using finger-prick devices or by wearing continuous monitoring sensors, which some patients consider as painful and burdensome.

The doctor also informs you that they are trialling a new method of monitoring without using finger prick devices, which involves microwave signals.

a) How much do you understand about how technology based on microwaves works? [tick one]

- Never heard of them
- Have heard but don’t understand
- Have some understanding
- Understand quite well
- Understand and could explain to others.

a) Are you aware that technology based on microwaves is used in healthcare?

- Yes18
- No

b) How do you feel about the use of technology based on microwaves in healthcare?

________________________________________________________________________________________________________________________________________________________________________________________________________________________________________________________________________________________________________________________________________

**Information 1: Why?**

- People with diabetes are required to check their blood sugar level 5 to 10 times a day by finger-prick tests.
- This could be perceived as intrusive to everyday life and painful.
- You could miss out on clinically important high and low blood sugar episodes if you forget to carry out a test.

c) After reading this information about the case study, I would accept the use of this technology to monitor my disease activity.

- Strongly agree
- Agree
- Unsure
- Disagree
- Strongly disagree

**Information 2: Benefits**

- This is a pain-free technique as there is no needle on the device. You just need to attach a small patch sensor on your arm.
- Monitoring glucose by microwaves provides continuous blood sugar level, non-invasively.
- By producing continuous information, you will get a trend of your blood sugar level across the whole day.
- You can enjoy your daily life without worrying about any interruption for finger-pricking.

d) After reading this information about the case study, I would accept the use of this technology to monitor my disease activity.

- Strongly agree
- Agree
- Unsure
- Disagree
- Strongly disagree

**Information 3: Risks**

- This sensor uses very weak microwaves to monitor your blood sugar level.
- Research is ongoing about the long-term effects of microwaves on human tissue.

e) After reading this information about the case study, I would accept the use of this technology to monitor my disease activity.

- Strongly agree
- Agree
- Unsure
- Disagree
- Strongly disagree

f) **If you are still unsure about whether you would accept the use of this technology, what other information would help you make a decision?**

- Research studies in humans
- Successful case studies of other patients
- Demonstration of how it works

Other. Please state: __________________________________________________________________________________________________________________________________________

l) **If you are still unsure about whether you would accept the use of this technology, what other information would help you make a decision?**

______________________________________________________________________________________________________________________________________________________________

g) If you would still not use the technology, why not?

______________________________________________________________________________________________________________________________________________________________

h) How would you like to receive the above information? [tick as many as apply]

- Leaflet
- Visual demonstration
- Verbal conversation with healthcare professional
- Website
- Other: ________________________

i) Who should first tell you about monitoring your health with technology based on microwave signalling?

__________________________________________________________________________________

**You will now read case studies of new technologies being developed to monitor specific medical conditions. Please imagine you are one of the patients.**

Case study 4: Patient Reported Monitoring of Rheumatoid Arthritis – An **A**pp for self-monitoring

You have been told by your doctor that you have Rheumatoid Arthritis. The doctor provides you with a course of treatment. They also tell you that there is a free **A**pp to monitor and measure your disease activity (including symptom severities and your ability to undertake various daily activities), with data shared to your clinical team.

a) How much do you understand about how mobile Apps work? [tick one]

- Never heard of them
- Have heard but don’t understand
- Have some understanding
- Understand quite well
- Understand and could explain to others.

b) Are you aware that mobile apps are used in healthcare?

- Yes
- No

c) How do you feel about mobile apps used in healthcare?

________________________________________________________________________________________________________________________________________________________________________________________________________________________________________________________________________________________________________________________________________

**Information 1: Why?**

- *Between appointments, your doctor has no idea how you are reacting to your new treatment.*
- *This app provides measurements of your disease activity over time to your clinical team.*
- *To do this you are required to complete a small number of questions as often as you feel necessary.*
- *You can use the app on any device including PC/laptop, tablet or phone*

d) After reading this information about the case study, I would accept to use mobile apps to monitor my disease activity.

- Strongly agree
- Agree
- Unsure
- Disagree
- Strongly disagree

**Information 2: Benefits**

*Appointments will be tailored to your needs.*

e) After reading this information about the case study, I would accept to use mobile apps to monitor my disease activity.

- Strongly agree
- Agree
- Unsure
- Disagree
- Strongly disagree

**Information 3: Risks**

- *Risks are minimal.*
- *You will be required to share your data (these measurements) with your clinical team.*

f) After reading this information about the case study, I would accept to use mobile apps to monitor my disease activity.

- Strongly agree
- Agree
- Unsure
- Disagree
- Strongly disagree

j) **If you are still unsure about whether you would accept the use of this technology, what other information would help you make a decision?**

- Research studies in humans
- Successful case studies of other patients
- Demonstration of how it works

Other. Please state: __________________________________________________________________________________________________________________________________________

g) If you would still not use the technology, why not?

______________________________________________________________________________________________________________________________________________________________

h) How would you like to receive the above information? [tick as many as apply]

- Leaflet
- Visual demonstration
- Verbal conversation with healthcare professional
- Website
- Other: ________________________

i) Who should first tell you about monitoring your health with mobile apps?

__________________________________________________________________________________

#### Section 5 – Sociodemographic information

Everyone taking part in this questionnaire is being asked to complete these questions, so that we can compare responses between different groups of people and see how representative our study is compared to the UK population as a whole. If you prefer not to answer any/all of these questions, please tick the box “prefer not to give a response”.

1. What is your age?

- Please specify __________
- Prefer not to answer

2. What is your sex?

- Female
- Male
- Other
- Prefer not to answer

3. What is your ethnic group? [select one]
A. White

- British
- Irish
- Other white background

B. Mixed

- White and Black Caribbean
- White and Black African
- White and Asian
- Other Mixed background

C. Asian or Asian British

- Indian
- Pakistani
- Bangladeshi
- Other Asian background

D. Black or Black British

- Caribbean
- African
- Other Black background

E. Chinese or other ethnic group

- Chinese
- Any other

F. I would prefer not to answer

- Prefer not to answer

4. What is your current employment status? [select one]

- In active paid work
- Retired
- Unemployed and seeking work
- Unemployed due to illness or disability
- Other, please specify: _________________
- Prefer not to answer

5. What is your highest educational qualification? [select one]

- No formal qualification
- O level/CSE/GCSE
- NVQ/vocational qualification
- A level
- First degree (e.g. BA, BSc)
- Higher degree (e.g. MSc, PhD)
- Other, please specify: ______________
- Prefer not to answer

6. What is your postcode? ________________
7. How is your health in general?

- Very good
- Good
- Fair
- Bad
- Very Bad
- Prefer not to answer

8. Do you have any existing health condition?

- No condition
- Yes. Prefer not to disclose.
- Deafness or partial hearing loss.
- Blindness or partial sight loss.
- Learning disability (for example, Down’s Syndrome)
- Learning difficulty (for example, dyslexia)
- Developmental disorder (for example, Autistic Spectrum Disorder or Asperger’s Syndrome)
- Mental health condition
- Physical disability
- Long-term illness, disease or condition
- Other condition. Please specify: _______________

Thank you for completing the survey.
