## Supplementary Material 2 for "Public attitudes towards the use of novel technologies in their future healthcare: A UK survey"

**Supplemental Material 2 – Demographic characteristics of respondents**

|  |  | **Survey** | | |
| --- | --- | --- | --- | --- |
|  |  | **n** | **%/Mean\|SD** | |
| Current age | | 1450 | 46.4 | 17.13 |
| Age Band | 18-24 | 214 | 14.8 | |
|  | 25-34 | 245 | 16.9 | |
|  | 35-44 | 219 | 15.1 | |
|  | 45-54 | 242 | 16.7 | |
|  | 55-64 | 211 | 14.6 | |
|  | 65-74 | 319 | 22.0 | |
| Gender | Female | 733 | 50.6 | |
|  | Male | 716 | 49.4 | |
|  | Other | 1 | 0.1 | |
|  | Prefer not to answer | 0 | 0.0 | |
| Ethnic group | British | 1162 | 80.1 | |
|  | Irish | 27 | 1.9 | |
|  | Other white background | 64 | 4.4 | |
|  | White and Black Caribbean | 8 | 0.6 | |
|  | White and Black African | 6 | 0.4 | |
|  | White and Asian | 17 | 1.2 | |
|  | Other Mixed background | 7 | 0.5 | |
|  | Indian | 19 | 1.3 | |
|  | Pakistani | 9 | 0.6 | |
|  | Bangladeshi | 5 | 0.3 | |
|  | Other Asian background | 13 | 0.9 | |
|  | Caribbean | 8 | 0.6 | |
|  | African | 15 | 1.0 | |
|  | Other Black background | 3 | 0.2 | |
|  | Chinese | 17 | 1.2 | |
|  | Any other | 5 | 0.3 | |
|  | Prefer not to answer | 65 | 4.5 | |
| Current employment status | In active paid work | 818 | 56.4 | |
|  | Retired | 307 | 21.2 | |
|  | Unemployed and seeking work | 80 | 5.5 | |
|  | Unemployed due to illness or disability | 53 | 3.7 | |
|  | Other, please specify: | 137 | 9.4 | |
|  | Prefer not to answer | 55 | 3.8 | |
| Highest educational qualification | No formal qualification | 54 | 3.7 | |
|  | O level/CSE/GCSE | 234 | 16.1 | |
|  | NVQ/vocational qualification | 167 | 11.5 | |
|  | A level | 295 | 20.3 | |
|  | First degree (e.g. BA, BSc) | 465 | 32.1 | |
|  | Higher degree (e.g. MSc, PhD) | 178 | 12.3 | |
|  | Other, please specify: | 28 | 1.9 | |
|  | Prefer not to answer | 29 | 2.0 | |
| Region | East | 129 | 8.9 | |
|  | East Midlands | 106 | 7.3 | |
|  | London | 192 | 13.2 | |
|  | North East | 56 | 3.9 | |
|  | North West | 160 | 11.0 | |
|  | South East | 202 | 13.9 | |
|  | South West | 142 | 9.8 | |
|  | West Midlands | 118 | 8.1 | |
|  | Yorkshire and Humberside | 121 | 8.3 | |
|  | Northern Ireland | 40 | 2.8 | |
|  | Scotland | 116 | 8.0 | |
|  | Wales | 68 | 4.7 | |
| Health in general | Very good | 246 | 17.0 | |
|  | Good | 830 | 57.2 | |
|  | Fair | 322 | 22.2 | |
|  | Bad | 50 | 3.4 | |
|  | Very Bad | 2 | 0.1 | |
|  | Prefer not to answer | 0 | 0.0 | |
| Health conditions | No condition | 926 | 63.9 | |
|  | Yes. Prefer not to disclose. | 29 | 2.0 | |
|  | Deafness or partial hearing loss. | 35 | 2.4 | |
|  | Blindness or partial sight loss. | 13 | 0.9 | |
|  | Learning disability (for example, Down's Syndrome) | 0 | 0.0 | |
|  | Learning difficulty (for example, dyslexia) | 10 | 0.7 | |
|  | Developmental disorder (for example, Autistic Spectrum Disorder or | 2 | 0.1 | |
|  | Asperger's Syndrome) | 5 | 0.3 | |
|  | Mental health condition | 68 | 4.7 | |
|  | Physical disability | 43 | 3.0 | |
|  | Long-term illness, disease or condition | 209 | 14.4 | |
|  | Other condition. Please specify: | 110 | 7.6 | |
| Index of Multiple Deprivation Quintile* | Most deprived: Quintile 1 | 233 | 18.9 | |
|  | Quintile 2 | 260 | 21.1 | |
|  | Quintile 3 | 283 | 23.0 | |
|  | Quintile 4 | 214 | 17.4 | |
|  | Least deprived: Quintile 5 | 243 | 19.7 | |
| Are you a frequent user of technology in your everyday life? | Yes | 1198 | 82.6 | |
|  | No | 252 | 17.4 | |
| Have novel technologies been used in the management of your health? | Yes | 137 | 9.4 | |
|  | No | 1140 | 78.6 | |
|  | Unknown | 173 | 11.9 | |
| How often do you have contact with these novel technologies for the management of your health? | Never | 1225 | 84.5 | |
|  | Once every few years | 61 | 4.2 | |
|  | Once a year | 33 | 2.3 | |
|  | Every few months | 57 | 3.9 | |
|  | Every month | 14 | 1.0 | |
|  | Every week | 22 | 1.5 | |
|  | Every day | 38 | 2.6 | |

*Index of Multiple Deprivation Quintile was designated from the postcode provided in the survey. For England, <https://imd-by-postcode.opendatacommunities.org/imd/2019>; for Wales, <https://statswales.gov.wales/Catalogue/Community-Safety-and-Social-Inclusion/Welsh-Index-of-Multiple-Deprivation>; for Scotland <https://nhsnss.org/services/practitioner/dental/scottish-index-of-multiple-deprivation-simd/>; for Northern Ireland <https://deprivation.nisra.gov.uk/>
