## Supplementary Material 3 for "Public attitudes towards the use of novel technologies in their future healthcare: A UK survey"

**Supplemental Material 3 - Acceptance of NHTs in future care according to respondents’ individual characteristics**

A single agreement score was calculated for each case study by taking the average agreement across 3 items that followed information explaining:

1. why the NHT should be used;
2. the benefits of the NHT;
3. the risks of the NHT.

In all cases, this was only calculated where all 3 items were answered. 1 was assigned to Strong Agree through to 5 for Strongly Disagree, so lower scores here indicated higher levels of agreement.

|  |  | Early detection of osteoarthritis | | | | | Virtual reality to support routine treatment | | | | Continuous glucose monitor for diabetes | | | | Patient Reported Monitoring of Rheumatoid Arthritis | | | |
| --- | --- | --- | --- | --- | --- | --- | --- | --- | --- | --- | --- | --- | --- | --- | --- | --- | --- | --- |
|  |  | **n** | **Med** | **IQR** | | **n** | | **Med** | **IQR** | | **n** | **Med** | **IQR** | | **n** | **Med** | **IQR** | |
|  |  |  |  | **LB** | **UB** |  |  |  | **LB** | **UB** |  |  | **LB** | **LB** |  |  | **LB** | **UB** |
| Age Band | 18-24 | 168 | 2.0 | 1.7 | 2.3 | 165 | | 2.0 | 2.0 | 2.3 | 165 | 2.0 | 1.7 | 2.3 | 165 | 2.0 | 1.3 | 2.0 |
|  | 25-34 | 203 | 1.7 | 1.3 | 2.3 | 202 | | 2.0 | 1.7 | 2.7 | 203 | 2.0 | 1.3 | 2.3 | 199 | 1.3 | 1.0 | 2.0 |
|  | 35-44 | 166 | 1.7 | 1.0 | 2.0 | 162 | | 2.0 | 1.3 | 3.0 | 167 | 2.0 | 1.3 | 2.7 | 158 | 1.3 | 1.0 | 2.0 |
|  | 45-54 | 190 | 1.7 | 1.3 | 2.0 | 177 | | 2.0 | 1.3 | 3.0 | 188 | 2.0 | 1.3 | 2.7 | 182 | 1.3 | 1.0 | 2.0 |
|  | 55-64 | 169 | 1.3 | 1.0 | 2.0 | 157 | | 2.3 | 1.3 | 2.0 | 172 | 2.0 | 1.3 | 2.7 | 170 | 1.3 | 1.0 | 2.0 |
|  | 65-74 | 243 | 1.3 | 1.0 | 2.0 | 226 | | 2.0 | 1.7 | 3.0 | 224 | 2.0 | 1.0 | 2.3 | 219 | 1.3 | 1.0 | 2.0 |
| Gender | Female | 589 | 1.7 | 1.3 | 2.0 | 573 | | 2.0 | 1.3 | 3.0 | 589 | 2.0 | 1.3 | 2.7 | 577 | 1.3 | 1.0 | 2.0 |
|  | Male | 549 | 1.7 | 1.3 | 2.0 | 515 | | 2.0 | 1.3 | 2.7 | 529 | 2.0 | 1.3 | 2.3 | 515 | 1.7 | 1.0 | 2.0 |
| Index of Multiple Deprivation Quintile | Most deprived: Quintile 1 | 182 | 1.7 | 1.3 | 2.3 | 178 | | 2.0 | 1.7 | 3.0 | 181 | 2.0 | 1.3 | 2.7 | 181 | 1.3 | 1.0 | 2.0 |
|  | Quintile 2 | 215 | 1.7 | 1.3 | 2.0 | 197 | | 2.3 | 1.3 | 3.0 | 211 | 2.0 | 1.3 | 2.3 | 198 | 1.7 | 1.0 | 2.0 |
|  | Quintile 3 | 224 | 1.7 | 1.3 | 2.0 | 209 | | 2.0 | 1.3 | 3.0 | 214 | 2.0 | 1.3 | 3.0 | 211 | 1.3 | 1.0 | 2.0 |
|  | Quintile 4 | 171 | 1.7 | 1.3 | 2.0 | 163 | | 2.0 | 1.3 | 3.0 | 169 | 2.0 | 1.3 | 2.3 | 162 | 1.0 | 1.0 | 2.0 |
|  | Least deprived: Quintile 5 | 186 | 1.3 | 1.0 | 2.0 | 181 | | 2.0 | 1.3 | 3.0 | 185 | 1.7 | 1.0 | 2.3 | 182 | 1.0 | 1.0 | 2.0 |
| Highest educational qualification | No formal qualification | 38 | 2.0 | 1.3 | 2.3 | 39 | | 2.3 | 2.0 | 3.3 | 42 | 2.0 | 1.3 | 3.0 | 37 | 2.0 | 1.3 | 2.7 |
|  | O level/CSE/GCSE | 172 | 1.7 | 1.3 | 2.0 | 170 | | 2.3 | 1.7 | 3.0 | 175 | 2.0 | 1.3 | 3.0 | 171 | 2.0 | 1.0 | 2.0 |
|  | NVQ/vocational qualification | 123 | 1.3 | 1.3 | 2.0 | 115 | | 2.3 | 1.7 | 3.0 | 124 | 2.0 | 1.3 | 2.7 | 119 | 1.7 | 1.0 | 2.0 |
|  | A level | 250 | 1.7 | 1.3 | 2.0 | 231 | | 2.0 | 1.3 | 2.7 | 240 | 2.0 | 1.3 | 2.3 | 233 | 1.3 | 1.0 | 2.0 |
|  | First degree (e.g. BA, BSc) | 384 | 1.7 | 1.3 | 2.0 | 372 | | 2.0 | 1.3 | 2.7 | 372 | 2.0 | 1.3 | 2.3 | 378 | 1.7 | 1.0 | 2.0 |
|  | Higher degree (e.g. MSc, PhD) | 141 | 1.7 | 1.3 | 2.0 | 134 | | 2.0 | 1.3 | 2.7 | 138 | 2.0 | 1.3 | 2.3 | 126 | 1.3 | 1.0 | 2.0 |
| Health conditions | No | 710 | 1.7 | 1.3 | 2.0 | 686 | | 2.0 | 1.3 | 2.7 | 701 | 2.0 | 1.3 | 2.3 | 691 | 1.7 | 1.0 | 2.0 |
|  | Yes | 429 | 1.3 | 1.3 | 2.0 | 403 | | 2.0 | 1.3 | 3.0 | 418 | 2.0 | 1.3 | 2.3 | 402 | 1.3 | 1.0 | 2.0 |
| Health in general | Very good | 195 | 1.3 | 1.0 | 2.0 | 186 | | 2.0 | 1.3 | 2.7 | 187 | 1.7 | 1.3 | 2.3 | 179 | 1.0 | 1.0 | 2.0 |
|  | Good | 642 | 1.7 | 1.3 | 2.0 | 627 | | 2.0 | 1.7 | 3.0 | 637 | 2.0 | 1.3 | 2.3 | 630 | 1.7 | 1.0 | 2.0 |
|  | Fair | 262 | 1.7 | 1.3 | 2.3 | 244 | | 2.3 | 1.7 | 3.0 | 257 | 2.0 | 1.3 | 2.7 | 249 | 1.7 | 1.0 | 2.0 |
|  | Bad or Very Bad | 40 | 1.3 | 1.0 | 2.0 | 32 | | 2.0 | 1.3 | 2.3 | 38 | 2.0 | 1.0 | 3.0 | 35 | 1.0 | 1.0 | 2.0 |
| Ethnic group | White British | 914 | 1.7 | 1.3 | 2.0 | 870 | | 2.0 | 1.3 | 3.0 | 899 | 2.0 | 1.3 | 2.3 | 877 | 1.3 | 1.0 | 2.0 |
|  | All other white | 66 | 1.7 | 1.3 | 2.0 | 62 | | 2.3 | 1.3 | 2.7 | 63 | 2.0 | 1.3 | 2.7 | 61 | 1.7 | 1.0 | 2.0 |
|  | Mixed/Multiple ethnic groups | 25 | 2.0 | 1.7 | 2.3 | 24 | | 2.7 | 1.8 | 3.2 | 23 | 2.0 | 1.3 | 2.7 | 23 | 1.3 | 1.0 | 2.0 |
|  | Asian/Asian British | 55 | 2.0 | 1.3 | 2.3 | 49 | | 2.0 | 1.7 | 2.3 | 54 | 2.0 | 1.7 | 2.7 | 51 | 1.3 | 1.0 | 2.0 |
|  | Black/African/Caribbean/Black British | 18 | 2.0 | 1.3 | 2.7 | 19 | | 2.0 | 1.3 | 2.3 | 18 | 2.2 | 1.7 | 3.0 | 18 | 2.0 | 1.7 | 2.7 |
|  | Other ethnic group | 4 | 2.2 | 1.5 | 2.7 | 4 | | 1.7 | 1.3 | 3.5 | 4 | 1.8 | 1.5 | 2.2 | 3 | 1.0 | 1.0 | 1.3 |
|  | Prefer not to answer | 57 | 2.0 | 2.0 | 2.0 | 61 | | 2.0 | 2.0 | 2.3 | 58 | 2.0 | 2.0 | 2.3 | 60 | 2.0 | 1.8 | 2.0 |
| Frequent user of technology | Yes | 960 | 1.7 | 1.3 | 2.0 | 919 | | 2.0 | 1.3 | 3.0 | 941 | 2.0 | 1.3 | 2.3 | 920 | 1.3 | 1.0 | 2.0 |
|  | No | 179 | 2.0 | 1.3 | 2.3 | 170 | | 2.3 | 2.0 | 3.0 | 178 | 2.0 | 1.7 | 3.0 | 173 | 2.0 | 1.0 | 2.3 |
| Novel technologies used in management of your health? | Yes | 110 | 1.7 | 1.3 | 2.0 | 106 | | 2.0 | 1.3 | 2.7 | 103 | 2.0 | 1.3 | 2.3 | 110 | 1.3 | 1.0 | 2.0 |
|  | No | 907 | 1.7 | 1.3 | 2.0 | 857 | | 2.0 | 1.3 | 3.0 | 885 | 2.0 | 1.3 | 2.3 | 858 | 1.3 | 1.0 | 2.0 |
|  | Unknown | 122 | 2.0 | 1.3 | 2.0 | 126 | | 2.0 | 1.7 | 2.3 | 131 | 2.0 | 1.7 | 2.3 | 125 | 2.0 | 1.0 | 2.0 |
| Contact with these novel technologies for the management of your health? | Never | 967 | 1.7 | 1.3 | 2.0 | 921 | | 2.0 | 1.3 | 3.0 | 952 | 2.0 | 1.3 | 2.3 | 920 | 1.7 | 1.0 | 2.0 |
|  | Once every few years | 51 | 1.7 | 1.3 | 2.3 | 52 | | 2.3 | 1.3 | 2.8 | 49 | 2.0 | 1.7 | 2.7 | 52 | 1.3 | 1.0 | 2.0 |
|  | Once a year | 23 | 2.0 | 1.3 | 2.3 | 23 | | 2.7 | 2.0 | 3.0 | 23 | 2.3 | 1.7 | 3.0 | 24 | 2.0 | 1.3 | 2.7 |
|  | Every few months | 36 | 2.0 | 1.3 | 2.2 | 35 | | 2.0 | 1.3 | 2.3 | 38 | 2.2 | 1.3 | 2.7 | 37 | 1.7 | 1.0 | 2.0 |
|  | Every month | 12 | 1.7 | 1.3 | 2.3 | 13 | | 2.7 | 2.0 | 3.0 | 12 | 2.0 | 1.5 | 2.7 | 12 | 1.3 | 1.0 | 2.0 |
|  | Every week | 15 | 1.7 | 1.0 | 2.0 | 14 | | 1.7 | 1.3 | 2.0 | 14 | 2.0 | 1.0 | 2.0 | 15 | 1.0 | 1.0 | 2.0 |
|  | Every day | 35 | 1.7 | 1.3 | 2.0 | 31 | | 2.3 | 2.0 | 3.0 | 31 | 1.7 | 1.0 | 2.0 | 33 | 1.3 | 1.0 | 2.0 |
| How much do you understand? (specific to case study) | Never heard of them | 318 | 2.0 | 1.3 | 2.3 | 113 | | 2.7 | 2.0 | 3.0 | 464 | 2.0 | 1.3 | 2.7 | 210 | 2.0 | 1.0 | 2.7 |
|  | Have heard but don't understand | 347 | 1.7 | 1.3 | 2.0 | 201 | | 2.3 | 2.0 | 3.0 | 272 | 2.0 | 1.3 | 2.7 | 123 | 2.0 | 1.0 | 2.3 |
|  | Have some understanding | 396 | 1.7 | 1.3 | 2.0 | 467 | | 2.0 | 1.7 | 3.0 | 263 | 2.0 | 1.3 | 2.3 | 305 | 2.0 | 1.0 | 2.0 |
|  | Understand quite well | 64 | 1.3 | 1.0 | 2.0 | 226 | | 2.0 | 1.3 | 2.3 | 96 | 1.7 | 1.0 | 2.0 | 267 | 1.3 | 1.0 | 2.0 |
|  | Understand and could explain to others. | 14 | 1.0 | 1.0 | 1.7 | 82 | | 1.3 | 1.0 | 2.3 | 24 | 1.0 | 1.0 | 1.5 | 188 | 1.0 | 1.0 | 1.3 |
| Are you aware of use? (specific to case study) | Yes | 748 | 1.7 | 1.3 | 2.0 | 206 | | 2.0 | 1.3 | 2.3 | 322 | 2.0 | 1.3 | 2.0 | 751 | 1.3 | 1.0 | 2.0 |
|  | No | 391 | 1.7 | 1.3 | 2.3 | 883 | | 2.0 | 1.7 | 3.0 | 797 | 2.0 | 1.3 | 2.7 | 342 | 2.0 | 1.0 | 2.3 |
